# Comparison of the antithrombotic strategy implemented to prevent early recurrence before closure of patent foramen ovale

**DOI:** 10.1101/2025.04.14.25325844

**Authors:** Marie Gachignard, François Derimay, Gilles Rioufol, Hélène Thibault, Muriel Rabilloud, Tae-Hee Cho, Laura Mechtouff, Laurent Derex, Elodie Ong, Julia Fontaine, Quentin Fleury, Pierre Pernot, Lucie Rascle, Paul Clottes

**Affiliations:** Department of vascular neurology, Hôpital Neurologique Pierre Wertheimer, Hospices Civils de Lyon, Bron, France; CarMeN, INSERM U1060, INRA U1397, INSA de Lyon, Université Lyon 1, Bron, France; Department of Interventional Cardiology, Cardiovascular Hospital, Hospices Civils de Lyon, France; Echocardiography Lab, Hôpital Cardiologique Louis Pradel, Hospices Civils de Lyon, France; Hospices Civils de Lyon, Pôle Santé Publique, Service de Biostatistique et Bioinformatique, Lyon, France; CNRS UMR 5558, Laboratoire de Biométrie et Biologie Évolutive, Équipe Biostatistique-Santé, Lyon, France

**Keywords:** Patent foramen ovale, early stroke recurrence, anticoagulant, antiplatelet, antithrombotic strategy, PFO closure

## Abstract

**Background:** Benefits of percutaneous patent foramen ovale (PFO) closure is demonstrated in PFO-associated stroke for secondary prevention, compared with medical treatment alone. Unfortunately, stroke recurrence can occur before PFO closure. However, no study has focused on the prevention of early cerebral ischemic recurrences that can occur before closure.

The aim of our study was to compare the antithrombotic strategy implemented to prevent early cerebral ischemic recurrences before PFO closure.

**Method:** This is a retrospective, single-center cohort study of adult patients with ischemic stroke who underwent PFO closure at Cardiology Hospital in Lyon between January 3, 2020, and November 22, 2023. The primary outcome was the occurrence of an ischemic recurrence, stroke or transient ischemic attack (TIA), before closure. Major bleeding represented the safety endpoint.

**Results:** In this retrospective cohort of 492 patients with an indication for PFO closure performed within one year following an ischemic stroke, 384 (78%) were under antiplatelet therapy (APT) and 108 (22%) under anticoagulant treatment (ACT). There were 15 early cerebral ischemic recurrences. All of these occurred under APT. Complete separation of the data prevented us to conclude with a logistic regression but suggested a significant link between APT and ischemic recurrence. No serious bleeding complication occurred.

**Conclusion:** Our retrospective cohort of PFO-related stroke patients suggests that early ischemic recurrences are more frequent with APT than with ACT, with no increase in hemorrhagic risk. The superiority of an antithrombotic strategy in this early time window (before PFO-closure) had not been previously studied, and our results need a randomized trial for confirmation.

## 1. Introduction

Ischemic stroke is a frequent and severe disease. Identifying the underlying etiology is an essential aspect of patient management, but approximately 25% of ischemic strokes remain cryptogenic. ^1^ In recent years, patent foramen ovale (PFO) has emerged from this category to become a recognized etiology of embolic stroke. ^2^ Several trials have demonstrated the benefits of percutaneous PFO closure for secondary prevention, compared with medical treatment alone. ^3^

Depending on the clinical situation, time to closure can range from a few days to several months. Unfortunately, stroke recurrence can occur before the closure of the PFO. To our knowledge, no study has focused on the prevention of these early cerebral ischemic recurrences before PFO closure. Some studies have compared long-term antithrombotic treatments for secondary prevention, with few results. ^4^ However, a recent meta-analysis has demonstrated a significant reduction in the risk of stroke recurrence with anticoagulant treatment (ACT) compared to antiplatelet therapy (APT), without evidence of an increased hemorrhagic risk. ^5^

Thus, medical practices are heterogeneous in this context and may potentially lead to more ischemic recurrences before closure, or more bleeding complications. As an example, the 2019 recommendations from the French Society of Neuro-Vascular (SFNV) and the French Society of Cardiology (SFC), specify that “anticoagulant therapy may be considered for the prevention of early recurrences while awaiting PFO closure, particularly in patients with a PFO and an ASA”, while reminding that “the benefit of anticoagulants compared to antiplatelet agents is unknown”. ^6^ The aim of our study was to compare the antithrombotic strategy used to prevent early cerebral ischemic recurrences before PFO closure.

## 2. Materials and methods

This is a retrospective, single-center cohort study of adult ischemic stroke patients who underwent PFO closure at Louis Pradel Cardiology Hospital in Lyon, France, between January 3, 2020, and November 22, 2023. Inclusion criteria were as follows: patients with ischemic stroke or transient ischemic attack (TIA) attributed to the PFO at the end of the etiological work-up, and for whom the indication for percutaneous PFO closure was confirmed and performed.

Patients who did not meet the characteristics of this population were excluded. Specifically, these were patients whose indication for PFO closure was non-neurological (as well as those for whom another etiology was identified retrospectively for the index event), and patients whose interatrial septum was confirmed to be completely closed during the procedure, which ruled out the possibility of the stroke was associated to a PFO. The lack of sufficient data regarding the antithrombotic therapy used was another exclusion criteria.

Additionally, we also decided to exclude patients with a delay of more than one year between the index event and the closure (this maximum delay was chosen as it corresponds to the 6-month period recommended by the SFNV, plus an additional 6-month period for extended cardiac rhythm monitoring if necessary) as well as patients who received both anticoagulant and antiplatelet therapy during the pre-PFO closure period.

Data were manually collected from the electronic medical records of patients.

The primary outcome was the occurrence of an ischemic recurrence, either a stroke or a TIA, before closure. Stroke was defined as clinical symptoms consistent with a stroke confirmed by MRI, or as an incidental finding on a brain MRI performed between the first ischemic event and the PFO closure. TIA was defined as suggestive transient clinical symptoms of neurovascular appearance with a normal brain MRI, with the diagnosis needing confirmation by a neurologist working in stroke unit.

The safety criteria was the occurrence of a severe hemorrhage, defined as intra- or extracerebral bleeding that led to hospitalization, disability or death.

All patients underwent transesophageal echocardiography (TEE) with bubble test (some on a pre-procedure table) to confirm PFO and to assess for ASA criteria (i.e. an abnormal, mobile aspect of the fossa ovale, with excursion into the left and/or right atrium of at least 10 mm).

Group comparisons were made using Fisher’s exact test for categorical variables and Mann-Whitney U-test for quantitative independent variables. Data are expressed as numbers and proportions (%) or median with interquartile range. We intended to build a logistic multivariate model that included every relevant variable (variables with a p-value < 0.2 in univariate analyses): type of antithrombotic treatment, age, initial ischemic event, ASA, delay to closure, diabetes and smoking. All statistical analyses were made using the Graph Pad Prism 9 software. A p-value <0.05 was considered significant.

The study was approved by the Scientific and Ethical Committee of Hospices Civils de Lyon (23-5443), no informed consent was required from participants.

## 3. Results

### Flow chart and population characteristics

A total of 738 patients were scheduled for a PFO closure during the period described. After application of the exclusion criteria, 492 patients were included in the analysis, including 108 (22%) patients in the anticoagulant group (ACT) and 384 patients (78%) in the antiplatelet group (APT). The flow chart is shown in figure 1.

**Figure 1.**
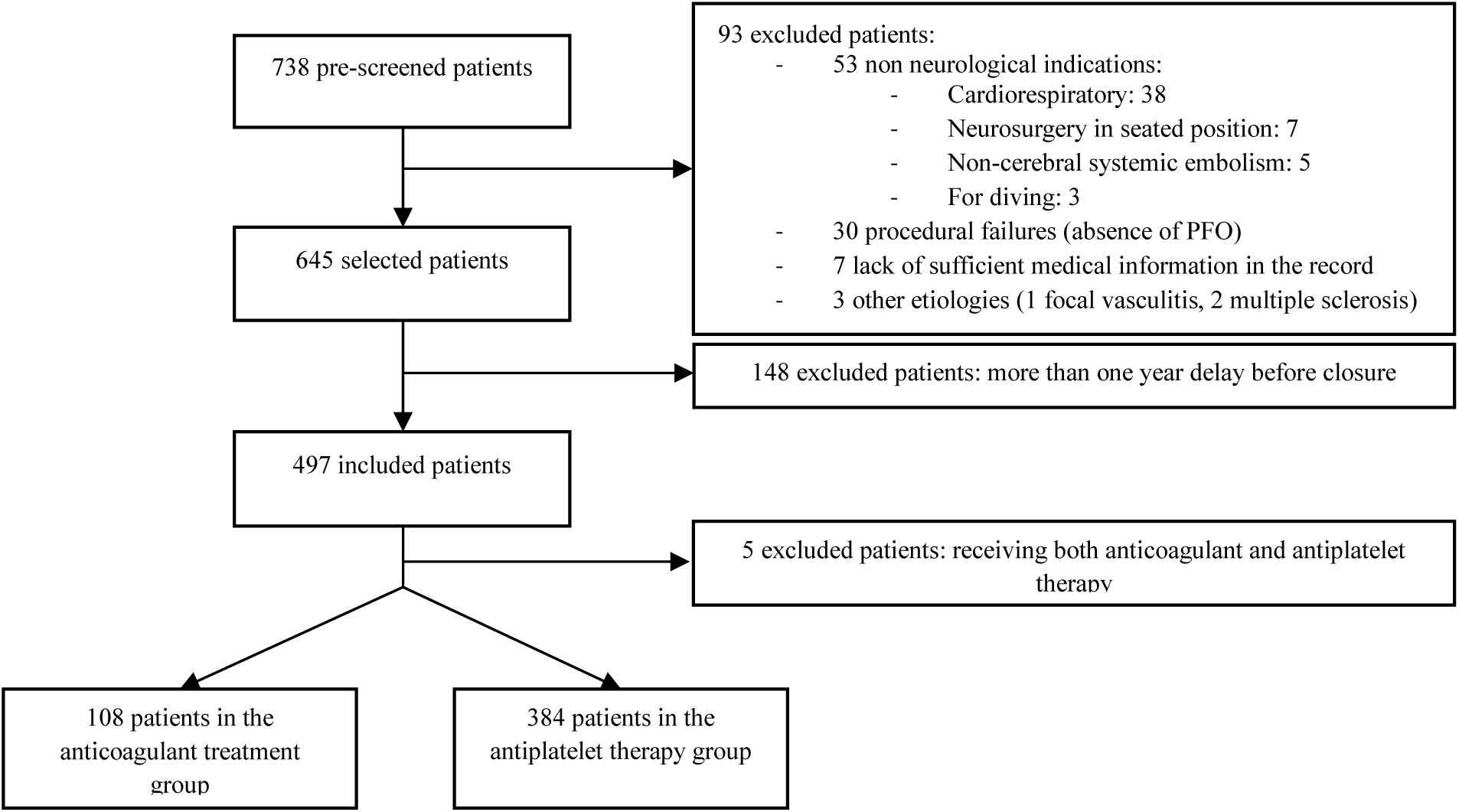
Flow chart

Among the 108 patients who received anticoagulation, 99 patients (91.6%) were on direct oral anticoagulants (DOACs), including 65 (60%) on Apixaban, 33 (30%) on Rivaroxaban, and 1 (0.9%) on Dabigatran. Three patients (2.8%) were on vitamin K antagonists (VKAs). The remaining 6 patients (5.6%) were treated with heparin (4 on Enoxaparin and 2 on Tinzaparin).

Among the 384 patients who received APT, 370 (96.4%) were on Aspirin (including 43 who received short-term dual antiplatelet therapy for 3 weeks in combination with Clopidogrel), and 14 (3.6%) were on Clopidogrel alone.

Table 1 summarizes the initial characteristics of the population and the comparison between the ACT and APT groups. The ACT group had significantly more ASA, a higher proportion of stroke than TIA as the index event, older patients, and a shorter time to closure compared to the APT group.

**Table 1.**
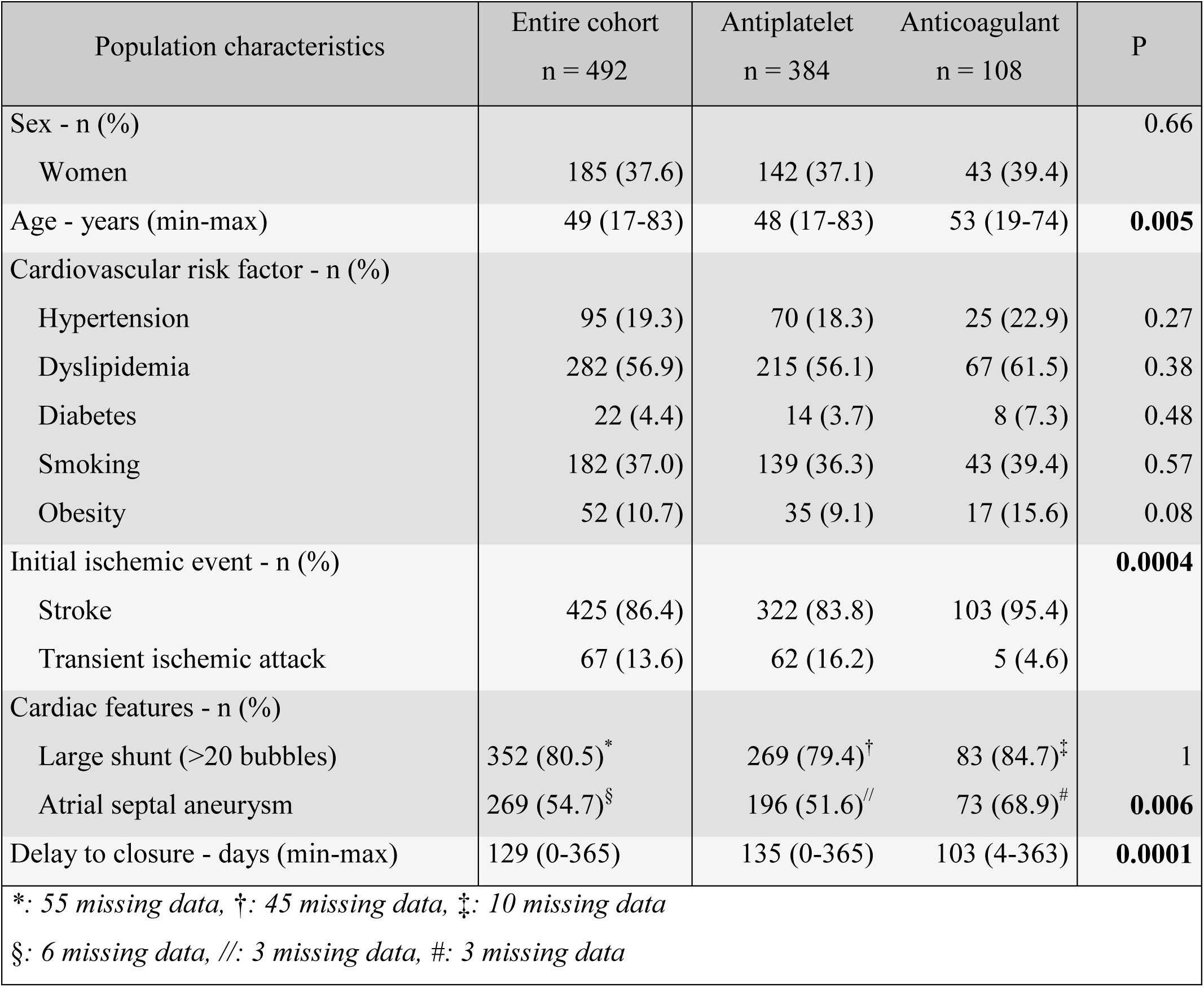
Cohort description.

### Early recurrences and hemorrhagic complications

There were 15 early recurrences, including 6 TIAs and 9 strokes. All occurred while on APT (1 on Clopidogrel, 2 on dual antiplatelet therapy with Aspirin and Clopidogrel, and the remaining 12 on Aspirin alone without previous dual antiplatelet therapy). The percentage of early recurrences among patients in the APT group was 3.9%.

Among the 9 early recurrent stroke patients, we had no information on the clinical manifestation for one, 2 were asymptomatic and discovered incidentally on brain MRI performed for an unknown reason, and 4 were associated with transient deficits. For the remaining 2 patients, the NIHSS increased by 2 points during the acute phase, and one of them underwent thrombolysis (mRS 2 at 6 months).

Of the 15 early recurrent ischemic event patients, 10 subsequently benefited from a change in anticoagulation therapy until closure (8 by DOAC and one by VKA), one from a change in antiplatelet class by Clopidogrel, one from an addition of Clopidogrel for 3 weeks, and no change was made for the remaining 3 patients.

Thirteen ischemic events (87%) occurred within 4 months of the initial event. Temporal curve of ischemic recurrences timing can be found in figure 2.

**Figure 2.**
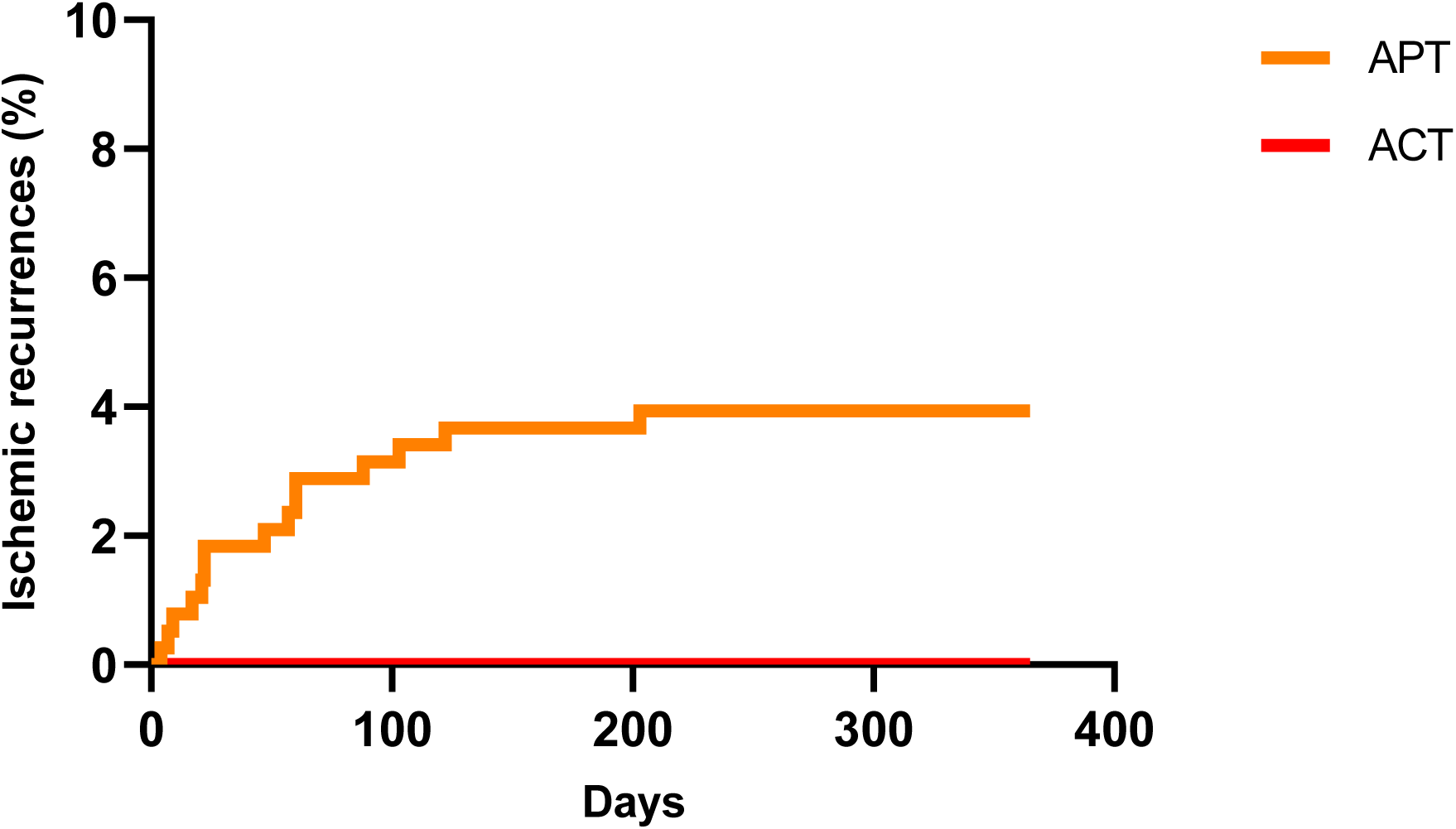
Temporal curve of ischemic recurrences timing

**Figure 3.**
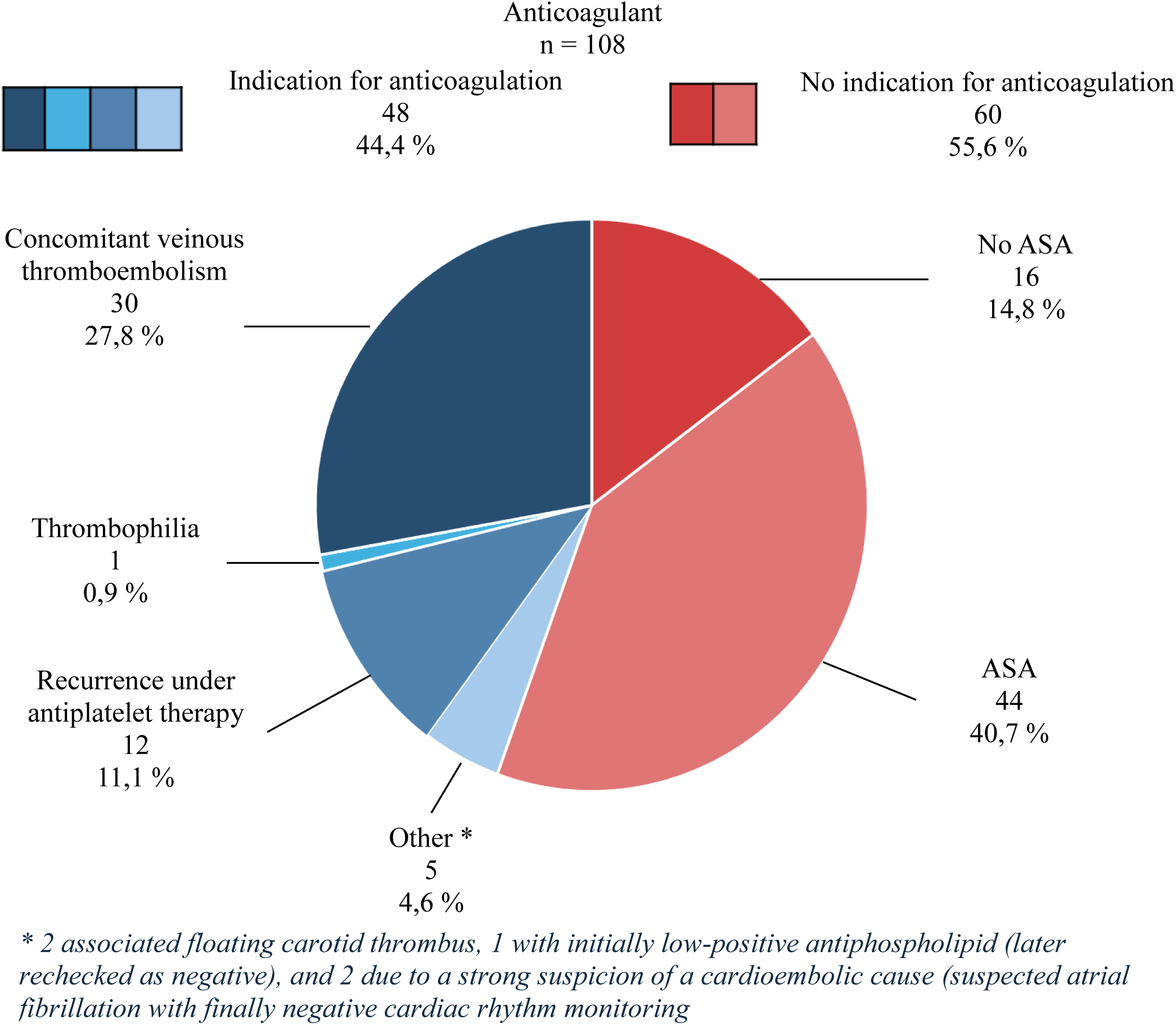
Reasons for anticoagulation

No major bleeding was reported in either group during the follow-up period.

Table 2 summarizes the characteristics of patients that experienced a recurrence. There was no significant difference between them and patients who did not experience a recurrence.

**Table 2.** Univariate comparison of characteristics between the “recurrence” and “no recurrence” groups.

| Population characteristics | No recurrence<br>n = 477 | Recurrence<br>n = 15 | P |
| --- | --- | --- | --- |
| Sex - n (%) |  |  |  |
| Women | 181 (38) | 4 (27) | 0.43 |
| Age - years (min-max) | 49 (17-83) | 48 (27-67) | 0.84 |
| Cardiovascular risk factor - n (%) |  |  |  |
| Hypertension | 94 (19.7) | 1 (6.7) | 0.32 |
| Dyslipidemia | 275 (57.7) | 7 (46.7) | 0.43 |
| Diabetes | 20 (4.2) | 2 (13.3) | 0.14 |
| Smoking | 178 (37.3) | 4 (26.7) | 0.19 |
| Obesity | 51 (10.7) | 1 (6.7) | > 0.999 |
| Initial ischemic event - n (%) |  |  | 0.71 |
| Stroke | 411 (86.4) | 14 (93.3) |  |
| Transient ischemic attack | 66 (13.6) | 1 (6.7) |  |
| Type of anti-thrombotic treatment - n (%) |  |  | 0.05 |
| Antiplatelet | 367 (77.4) | 15 (100) |  |
| Anticoagulant | 108 (22.6) | 0 (0) |  |
| Cardiac features - n (%) |  |  |  |
| Large shunt (>20 bubbles) * | 343 (80.5) † | 9 (60) ‡ | > 0.999 |
| Atrial septal aneurysm § | 260 (55.2) // | 9 (60) | 0.80 |
| Delay to closure - days (min-max) | 127 (0-365) | 135 (26-337) | 0.29 |
| *: 55 missing data, †: 51 missing data, ‡: 4 missing data, §: 6 missing data, //: 6 missing data |  |  |  |

As described in the methods, we intended to perform a logistic regression with “ischemic recurrence” as the outcome variable. However, the analysis was impossible to perform due to complete separation, meaning that one covariate perfectly predicted the outcome variable.

The variable responsible here is certainly “type of antithrombotic treatment”, as every ischemic recurrence occurred in the APT group. Therefore, a significant relationship between APT and ischemic recurrence can be strongly suspected, as under the assumption that both treatments have the same efficacy in preventing recurrence, the probability that all 15 patients who recurred belong to the same treatment group is 2.3×10^-3^. In this situation however, it is not possible to obtain a p value or odds ratio.

### Reason for anticoagulation

Among patients on ACT, we identified 48 (44.5%) for whom the indication was clear, as detailed in Figure 4. We found no obvious justification for ACT among the remaining 55.5% of patients on ACT. Among them, we identified 44 patients (40.7%) in whom the PFO was associated with an ASA, which is a debated indication for anticoagulation. There was no ASA for in the remaining 16 patients.

## 4. Discussion

In this retrospective cohort of 492 patients with a valid indication for PFO closure performed within one year following an ischemic stroke, there were 15 early cerebral ischemic recurrences. All occurred under APT. Complete separation prevents us to conclude with absolute certainty, but strongly suggests a significant association between APT and ischemic recurrence.

To the best of our knowledge, this is the first study to investigate antithrombotic strategy before PFO closure in stroke patients. The number of included patients is substantial, surpassing most previous cohort comparing these two types of treatment in the context of PFO. ^4,5,7^

The main result of our study aligns with recent data showing a possible reduction in the long-term risk of stroke recurrence with ACT compared to APT in PFO patient. ^5^ It is to note that most recurrences were clinically benign, including 6 TIA and 6 strokes with transient symptoms.

This result is also physiologically consistent with the main hypothesis regarding the mechanism of ischemic stroke originating from PFO: paradoxical venous embolism through the PFO ^8^, formation of an in situ thrombus, ^9^ and/or the formation of thrombi in the left atrium, secondary to left atrial cardiopathy induced by the PFO and exacerbated by ASA, which promotes both conduction disturbances and arrhythmias, without necessarily revealing atrial fibrillation. ^10,11^

The percentage of recurrences in the APT arm, 3.9% for the duration of the study, is consistent with the existing literature. ^12–14^ However, our follow-up time was of less than a year for most patients, with a median closure time of 4 months. Nonetheless, recurrences occured early in the follow-up period.

The absence of severe hemorrhagic complications contrasts with the results of previously published studies. ^5^ This result can be explained by two factors. First, the short duration of anticoagulation (less than one year, with a median of approximately 4 months). Second, unlike most previous comparative studies, a very large majority of our anticoagulated patients (92%) were treated with DOACs, including 60% with Apixaban. Their safety profile is superior to that of VKA as demonstrated for the prevention of atrial fibrillation ^15^, and a recent randomized trial of over 1,000 patients also found no significant increase in major bleeding risk with Apixaban compared to Aspirin (in patients with cryptogenic stroke and atrial cardiopathy). ^10^

Time to percutaneous closure was not associated with the risk of recurrence. This may be explained by the fact that most ischemic recurrences occurred early (within the first three months). Obviously, PFO closure should be performed as soon as possible after the PFO has been considered causal. However, availability of additional examinations (mainly cardiac monitoring) and of the closing procedure are two limitations that extend the time before closure. This unavoidable delay between stroke and closure, combined with the early recurrence of events, further increase the need for an optimization of the antithrombotic strategy before closure.

Anticoagulation practices were heterogeneous within our cohort, with 22% of patients receiving ACT. This percentage is slightly lower than the 30% of patients anticoagulated for secondary prevention in PFO-associated stroke (which, however, includes patients for whom closure was not indicated at the time, due to lack of evidence, or for whom it was contraindicated or refused). ^4,5^ Among the 55.5% of anticoagulated patients for whom the reason was not clearly established in the medical records, we estimated that the associated ASA in 40.7% probably led to this anticoagulation. On one hand, it is accepted that the association of ASA and PFO increases the risk of recurrence. ^16^ On the other hand, the expert consensus from the SFNV that discusses this issue may have been considered. ^6^ This hypothesis is supported by the fact that there was significantly more ASA in the ACT group than in the APT group.

The main limitation of this study is its retrospective nature based on the closure procedure, which may have, by definition, excluded patients who could have died or been ruled out of the closure procedure due to neurological or other complications, and non-randomized design. Multivariate analysis helps to account for differences between the two patient groups, but complete separation of the data prevents us to conclude with certainty on our primary outcome. Another significant limitation is its monocentric design. Indeed, we do not have visibility on practices at other French or international centers. Therefore, we chose a primary endpoint that combined the occurrence of constituted ischemic recurrences revealed by consistent symptoms, or incidental finding, with transient recurrences, to enhance the study’s power. These were diagnosed by experienced vascular neurologists, limiting the risk of differential diagnosis, and it is to note that there was no TIA in the ACT group.

## Conclusion

Our retrospective cohort of PFO-related stroke patients suggests that early ischemic recurrences are more frequent with APT than with ACT, with no increase in hemorrhagic risk. The superiority of an antithrombotic strategy in this early time window (before PFO-closure) had not been studied before, and our results will eventually need a randomized study for confirmation.

## Data Availability

The data supporting the findings of this study are available from the corresponding author upon reasonable request. All relevant data are included in the manuscript and its supplementary materials.

## Acknowledgments

none

## Sources of Funding

none

## Disclosures

none

## Notes

### Competing Interest Statement

The authors have declared no competing interest.

### Clinical Trial

It is a retrospective study.

### Funding Statement

No external funding was received for this work. The authors and their institutions did not receive any payment or services from a third party for any aspect of the submitted work.

